# High-Resolution Melt Curve Analysis: An Approach for Mutation Detection in the TPO Gene of Congenital Hypothyroid Patients in Bangladesh

**DOI:** 10.1101/2023.10.17.23297147

**Authors:** Mst. Noorjahan Begum, Rumana Mahtarin, Md Tarikul Islam, Nusrat Jahan Antora, Suprovath Kumar Sarker, Nusrat Sultana, Abu A. Sajib, Abul B.M.M.K Islam, Hurjahan Banu, M A Hasanat, Kohinoor Jahan Shyamaly, Suraiya Begum, Tasnia Kawsar Konika, Shahinur Haque, Mizanul Hasan, Sadia Sultana, Taufiqur Rahman Bhuiyan, Kaiissar Mannoor, Firdausi Qadri, Sharif Akhteruzzaman

## Abstract

Thyroid Peroxidase (*TPO*) is known to be the major gene involved in Congenital hypothyroid patients with thyroid dyshormonogenesis. This present study aimed to establish high-resolution melting (HRM) curve analysis as a supplementary mutation detection approach of Sanger sequencing targeting commonly found mutations c.1117G>T, c.1193G>C, and c.2173A>C in the TPO gene in Bangladeshi patients. We enrolled 36 confirmed cases of congenital hypothyroid patients with dyshormonogenesis to establish the HRM method. Blood samples were collected, and genomic DNA was isolated for molecular techniques. Among the 36 specimens, 20 were pre-sequenced, and mutations were characterized through Sanger sequencing. The pre-sequenced specimens (n=20) were then subjected to real-time PCR-HRM curve analysis to get the appropriate HRM condition capable of differentiating heterozygous and homozygous states for the three mutations from the wild-type state. Furthermore, 16 unknown specimens were subjected to HRM analysis to validate the method. This method showed 100 percent sensitivity and specificity to distinguish wild-type alleles from homozygous or heterozygous states (c.1117G>T, c.1193G>C, and c.2173A>C) of alleles commonly found in Bangladeshi patients. The HRM data was found to be similar to the sequencing result, thus confirming the validity of the HRM approach for *TPO* gene mutation.

In conclusion, the established HRM-based molecular technique targeting c.1117G>T, c.1193G>C, and c.2173A>C mutations could be used as a high throughput, rapid, reliable, and cost-effective screening approach for the detection of all common mutations in *TPO* gene in Bangladeshi patients with dyshormonogenesis.

## 1. Introduction

Congenital hypothyroidism (CH) is the endocrine disorder in which thyroid hormone deficiency occurs at birth and is the most common preventable cause of mental retardation [1-3]. This condition can lead to different clinical complications such as irreversible brain damage, delayed developmental milestones, lethargy, and slowdown of the body’s overall metabolism of the patients if untreated. Early detection and initiation of treatment can reverse such complications [4]. The CH frequency is 1 in 3000-4000 newborns worldwide, whereas it is much higher in Bangladesh [5-9]. 11 genes, including thyroid gland development and thyroid hormone biosynthesis, have been documented [10]. A defect in thyroid gland development due to mutations in both gene alleles of the pathway is known as thyroid dysgenesis. In contrast, a defect in thyroid hormone biosynthesis due to mutations in both alleles of a gene of the pathway is called thyroid dyshormonogenesis (TDH). TDH occurs due to mutations commonly found in seven genes, including TPO as a significant contributor to them [11, 12]. Since thyroid hormones are iodinated, TPO catalyzes the iodination steps, and mutations in the TPO gene may cause either total iodide organification defect (TIOD) or partial iodine organification defect (PIOD). Different countries conducted several studies on screening and identification of mutations in the TPO gene causing TIOD and PIOD [13-18]. Our previous study investigated that only genetic causes accounted for all of the CH patients with dyshormonogenesis in hospital settings in Bangladesh and didn’t find any other aetiology [19]. Since CH is easily treatable, it is important to investigate the aetiology, which would help to determine how long the patients need hormone replacement therapy. The CH patients with genetic aetiology need lifelong hormone therapy [20]. Sometimes, neonatal CH screening using biochemical tests becomes difficult due to the presence of maternal TSH in the specimens of neonates, and this problem can be overcome by genetic screening at an early age [20]. There is limited data on investigating the genetic causes of CH in Bangladesh. However, we have performed several genetic studies and found mutations in patients with thyroid dyshormonogensis [21] and thyroid dysgenesis [22].

High-resolution melting (HRM) curve analysis is one of the molecular tests, which is a high throughput real-time PCR technique based on the melting properties of double-stranded DNA. HRM can differentiate genetic variations such as homozygous or heterozygous states for specific mutations compared to the wild-type state in various genetic diseases, including autosomal recessive, autosomal dominant, and X-linked recessive disorders [23-26].

This study aimed to establish HRM curve analysis as a supplementary screening approach of Sanger sequencing targeting three common mutations in Bangladeshi patients. It could play an essential role in screening mass populations since it is faster, cheaper, and more reliable to detect genetic variations for improving the dimensions of newborn screening, which is neglected in Bangladeshi children.

## 2. Methods and Materials

### 2.1. Study participant enrolment and specimen collection

We enrolled a total of 36 confirmed cases of congenital hypothyroid children with dyshormonogenesis confirmed by an ultrasonogram of the thyroid gland in the clinical settings of the National Institute of Nuclear Medicine and Allied Sciences (NINMAS) and Department of Endocrinology, Bangabandhu Sheikh Mujib Medical University (BSMMU), Dhaka, Bangladesh.

### 2.2. Ethics approval and consent to participate

This study was approved by the Ethical Review Board for Human Studies of BSMMU and the Human Participants Committee, University of Dhaka (CP-4029) on 16 May 2017. After obtaining the ethical approval, we enrolled patients and collected their specimens from 1 June 2017 to 31 December 2019. Blood specimens were collected from the participants with informed written consent from their parents or guardians.

### 2.3. Laboratory investigation

#### 2.3.1. DNA isolation, PCR amplification, and Sanger Sequencing

Blood specimens (3 mL) were collected, and genomic DNA was isolated using the QIAGEN FlexiGene® DNA Kit, followed by PCR and Sequencing [19]. After that, 20 pre-sequenced specimens were subjected to HRM, and then 16 unknowns were tested.

#### 2.3.2. Method setup and validation of High-Resolution Melt curve analysis

For analysis of *TPO* gene mutations by HRM method, we targeted three nonsynonymous mutations in the *TPO* gene identified by Sanger sequencing. For this purpose, we designed three sets of primers for detecting c.1117G>T and c.1193G>C mutations in exon eight and c.2173A>C mutation in exon 12. The primer sequences are listed in Table 1. First of all, those mentioned above, 20 pre-sequenced specimens with known mutations were used as reference samples to set up the HRM method. Homozygous, heterozygous, and wild-type specimens for the specific mutations were subjected to HRM curve analysis. Finally, 16 unknown samples were run to validate the method. These 16 samples were further tested by Sanger sequencing to confirm the mutation and validate the HRM approach.

**Table 1:**
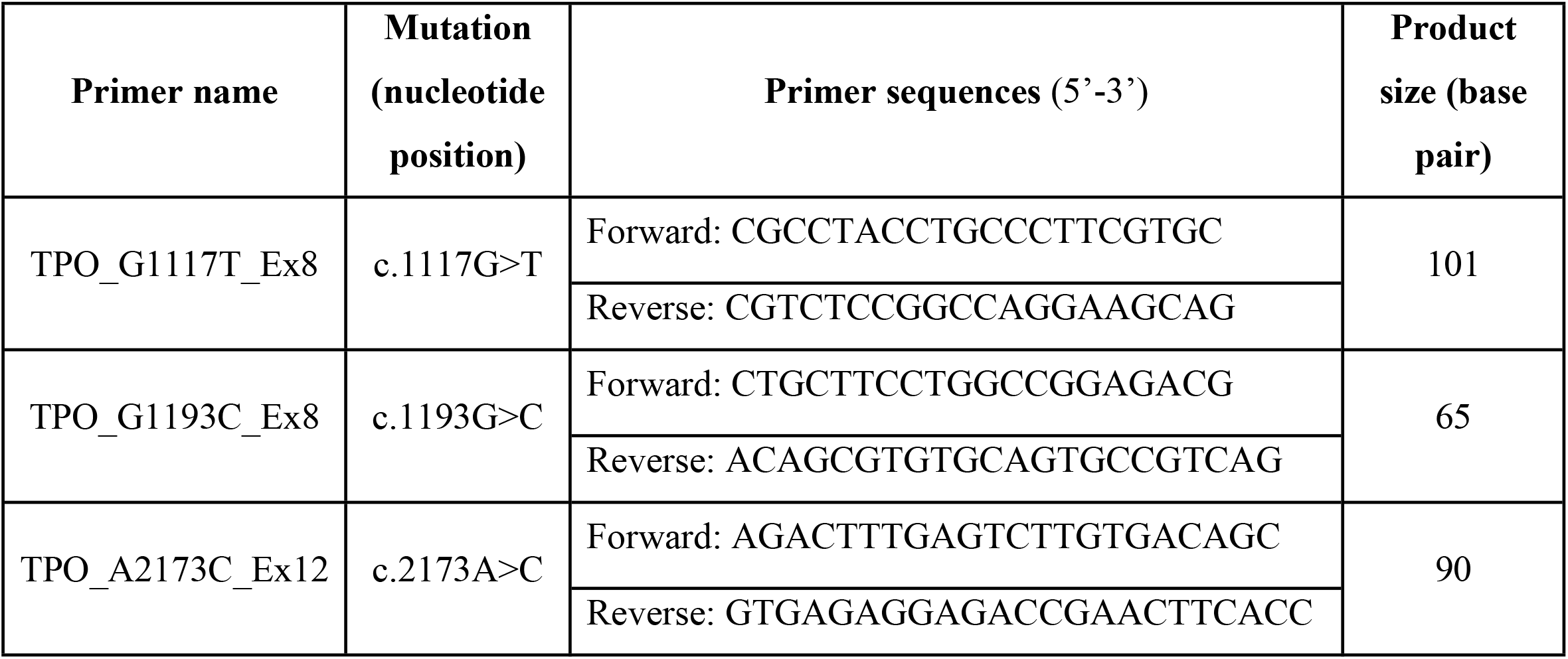
List of primers used in HRM curve analysis.

To amplify the target sequence, a master mix was prepared following the protocol provided with the Precision Melt Supermix kit (Bio-Rad). To a reaction mixture of 5 μL of 2X precision melt super mix, 0.2 μL of each forward and reverse primer and 1 μL of DNA (50 ng); 3.6 μL nuclease-free water were added to make the reaction volume up to 10 μL. Moreover, for detection of c.1193G>C mutation by HRM, 8mM MgCl_2_ was added to the reaction mixture, and the reaction volume was adjusted accordingly. The cyclic condition was divided into two steps, namely real-time PCR amplification followed by melt curve analysis using a single program. The real-time PCR-based HRM was performed on a CFX96 Touch™ Real-Time PCR machine (Bio-Rad). The real-time PCR cyclic condition was as follows: initial denaturation at 95°C for 3 min; 40 cycles of denaturation at 95°C for 10 s, annealing at 60°C for 15 s and extension at 72°C for 15 s. After completion of the real-time PCR, the subsequent melt curve program was initiated through cycles of denaturation at 95°C for 30 s, renaturation at 60°C for 1 min, and then melting at 65°C to 95°C with an increment of 0.1°C per 5 s for c.1117G>T and c.2173A>C mutations. And notably, an increment of 0.2°C per 5 s was used for c.1193G>C mutation. After completion of the real-time PCR-HRM, the data were analyzed using Precision Melt Analysis™ Software (BioRad). The melt curve shape sensitivity for cluster detection was set to 100%. The difference in the Tm threshold for the cluster detection was set to 0.1 to 0.2, and the normalized and temperature-shifted views were used for analysis. The results of pre-sequenced samples were compared with HRM analysis, and an additional 16 unknown samples were subjected to validate the HRM method using the same procedure that was followed for the pre-sequenced known samples. Finally, the normalized melt curve and the difference curves for both wild-type and mutant specimens (homozygous and heterozygous) were calculated and analyzed to detect the mutations in the *TPO* gene.

## 3. Results

Among 36 participants, 21 (58.33%) were males, and 15 (41.67%) were females. The average age of the participants was 7.97±4.29 (years) mean± SD, and the BMI was 17.0±4.4 (Kg/m^2^) mean± SD.

### 3.1. Screening of c.1117G>T mutation using HRM analysis

To establish a rapid HRM-based screening approach targeting the c.1117G>T variant, a pair of primers (TPO_G1117T_Ex8) was designed that flanked the c.1117G>T variant. Then, the pre-genotyped samples were subjected to real-time PCR followed by HRM analysis. The fluorescence started to drop quickly at the initial melting phase for heterozygous samples. However, in the later phase of melting, the homozygous samples started to lose fluorescence intensity quicker than the heterozygous samples, and as a consequence, they crossed each other at a certain point of melting, making them distinguishable from each other. Similar to the normalized melting curve (Fig 1), the temperature-shifted difference curve could generate distinctive melting patterns for the wild-type, homozygous and heterozygous specimens (Fig 2). The reliability of the method was further validated by analyzing 16 unknown samples with dyshormonogesis. At first, these samples were tested by HRM, and then Sanger sequencing was done to check the sensitivity and specificity of the method. A total of 5 specimens had c.1117G>T variant in heterozygous states, six specimens were in a homozygous state, and the remaining 5 had the wild type allele. The HRM result was consistent with the sequencing data, implying that sensitivity and specificity for detecting the c.1117G>T variant was 100% for both homozygous and heterozygous alleles.

**Fig 1.**
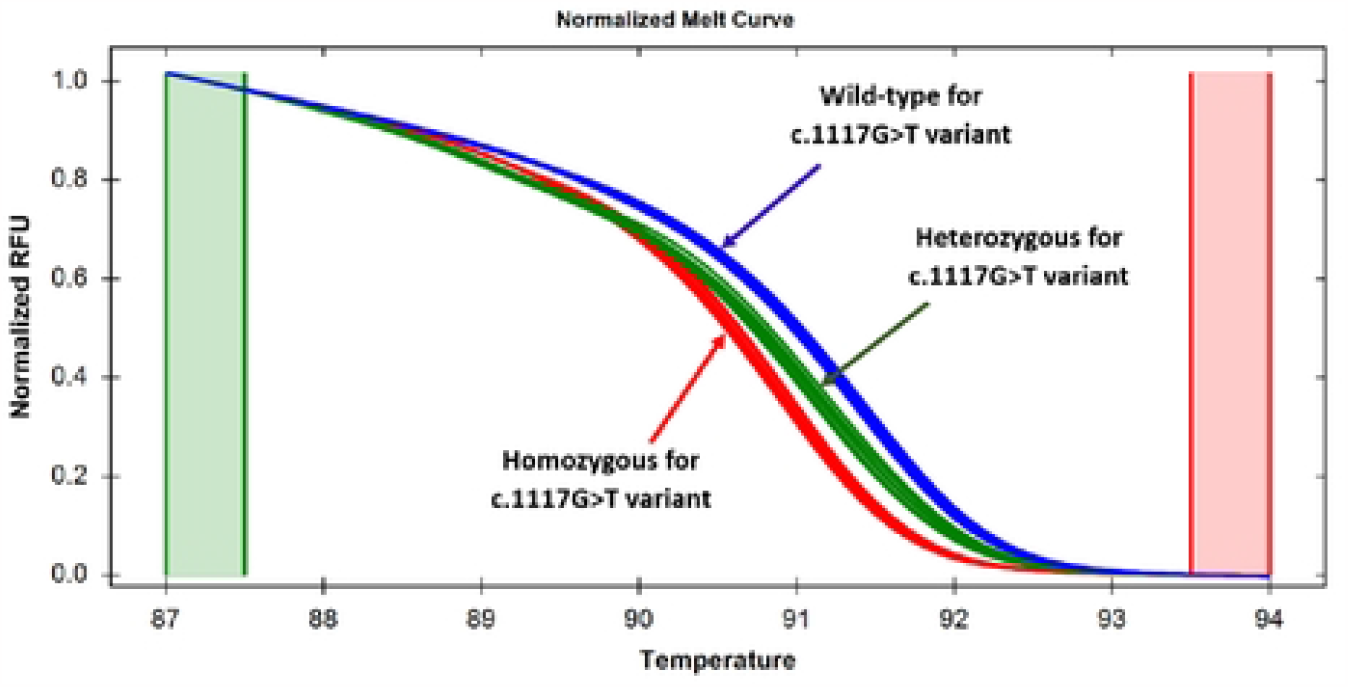
Normalized melt curves for the specimens targeting the c.1117G>T variant in exon-8. Normalized melt curves showing that the specimens with homozygous and heterozygous states are clearly distinguishable from the wild-type specimens, as manifested by the difference in relative fluorescence unit.

**Fig 2.**
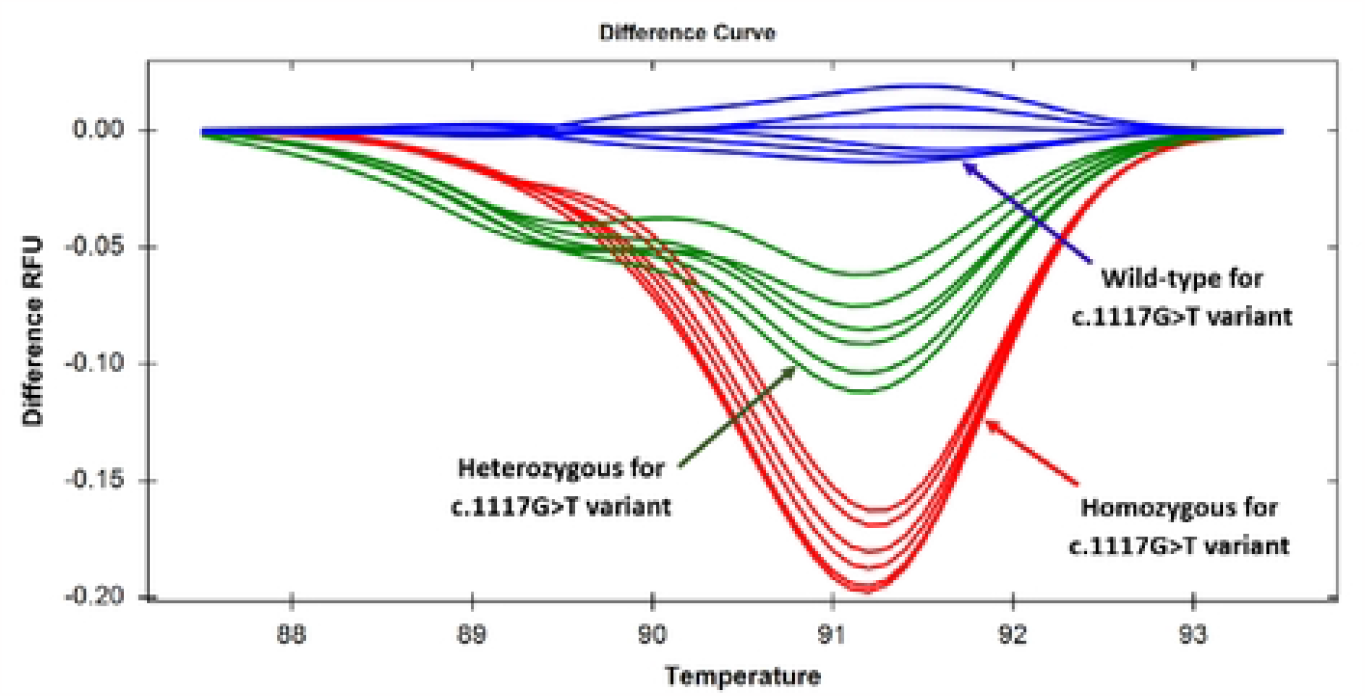
Difference curves generated by specimens targeting the c.1117G>T variant in exon-8. Discernable changes in three difference curves were showing that the specimens with homozygous and heterozygous states are clearly distinguishable from the wild type allele, as manifested by the difference in relative fluorescence unit.

### 3.2. Screening of c.1193G>C mutation using HRM analysis

The second set of primers, namely TPO_G1193C_Ex8, was used for the analysis of the c.1193G>C variant by the HRM approach. When the pre-genotyped samples were subjected to HRM analysis, three different clusters were observed in the melt curve analysis. One of the clusters corresponded to the heterozygous samples; the other two were for the homozygous samples and for the wild-type samples (Fig 3). However, in the difference curve analysis, three different clusters were clearly observed for homozygous, heterozygous, and wild-type variants of c.1193G>C (Fig 4). A similar observation was observed with 16 unknown samples that were subjected to HRM analysis. Four out of 16 samples came out as heterozygous by HRM analysis, and this result was consistent with the sequencing data. However, eight homozygous and four wild-type samples formed different clusters in this case. This observation implies that both heterozygous and homozygous states for c.1193G>C variant could be detected with 100% sensitivity and specificity.

**Fig 3.**
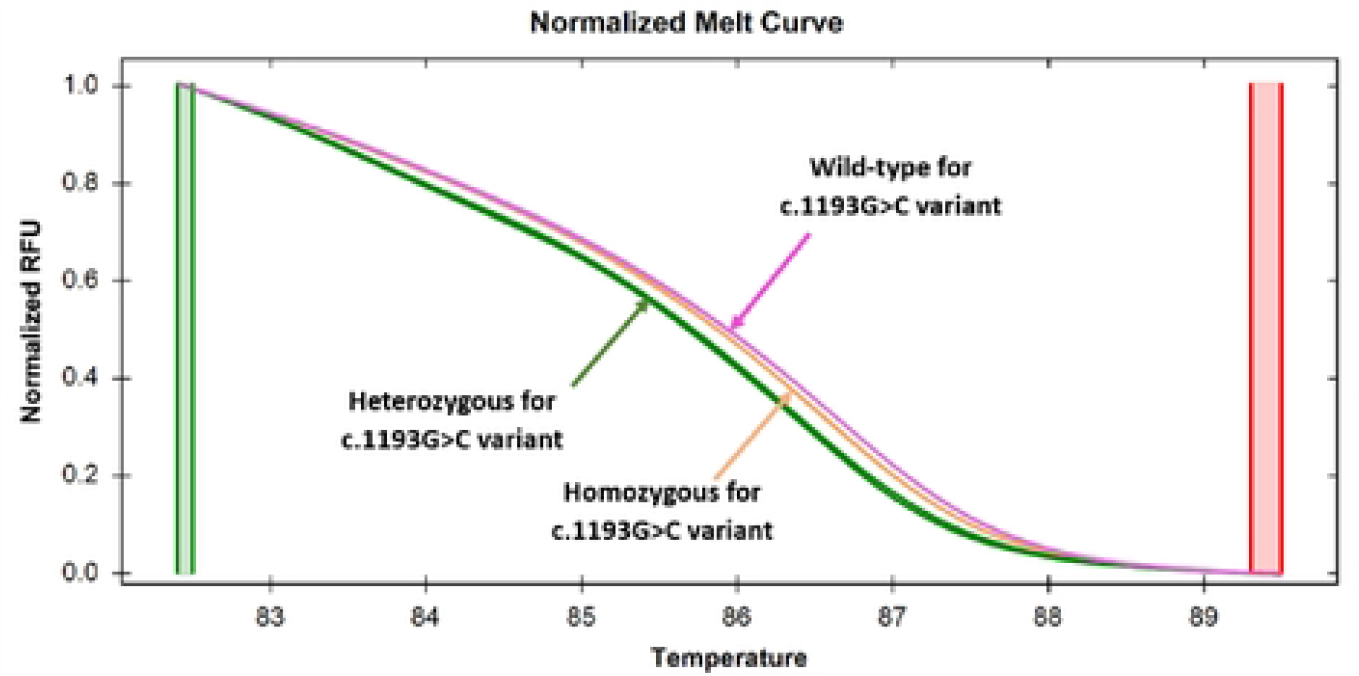
Normalized melt curves generated by specimens targeting the c.1193G>C variant in exon-8. Discernable changes in normalized melt curves were showing that the specimens with homozygous (orange color) and heterozygous (green color) states are clearly distinguishable from the wild type (purple color) alleles, as manifested by the difference in relative fluorescence unit.

**Fig 4.**
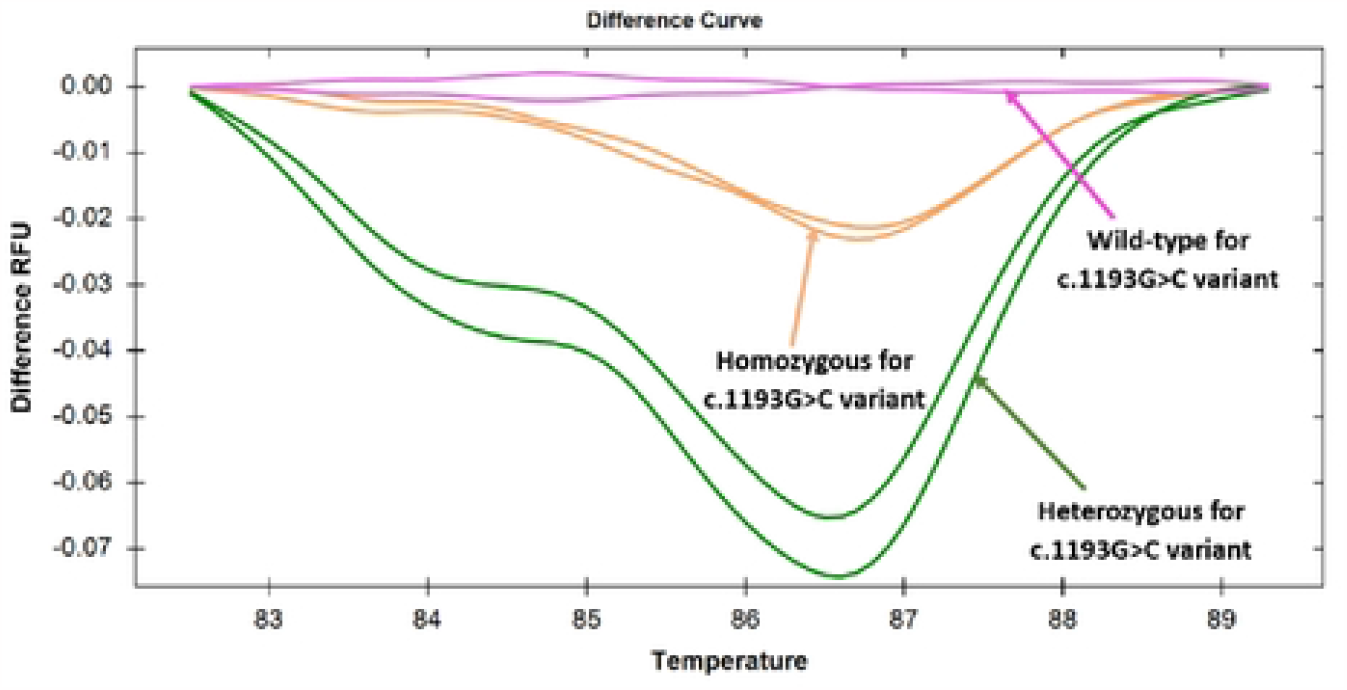
Difference curves for specimens targeting the c.1193G>C variant in exon-8. Discernable changes in difference curves were showing that the specimens with homozygous and heterozygous states are clearly distinguishable from the wild-type states, as manifested by the difference in relative fluorescence unit.

### 3.3. Screening of c.2173A>C mutation using HRM analysis

The third set of primers, namely TPO_A2173C_Ex12, was used to analyze another TPO gene mutation designated as c.2173A>C. The pre-genotyped wild type, homozygous c.2173A>C, and heterozygous c.2173A>C specimens were subjected to HRM analysis. The wild type, homozygous and heterozygous variants formed distinct clusters (Fig 5 and Fig 6). The homozygous c.2173A>C substitution resulted in an increase in melting temperature, and thus, the specimens with homozygous c.2173A>C variant had higher fluorescence intensity than that with the wild-type allele during melting, as manifested by the relative fluorescence unit (Fig 5). On the other hand, although the specimens with the heterozygous c.2173A>C variant followed a melting pattern with lower fluorescence intensity initially compared to the wild type, and the melting curve patterns almost overlapped with each other in a later stage (upper panel of Fig 5). Thus, homozygous c.2173A>C, heterozygous c.2173A>C, and the wild-type alleles were discernable from each other. Similar to the normalized melting curve, the difference curve analysis could also distinguish different states involving the c.2173A>C variant (Fig 6). HRM analysis of 16 unknown samples targeting the c.2173A>C variant was also 100% sensitive and specific. The HRM approach showed that 5 out of 16 unknown samples were wild type, 6 were homozygous, and the rest 5 were heterozygous. Sequencing of those 16 unknown samples revealed that the PCR HRM-based result was consistent with the sequencing result.

**Fig 5.**
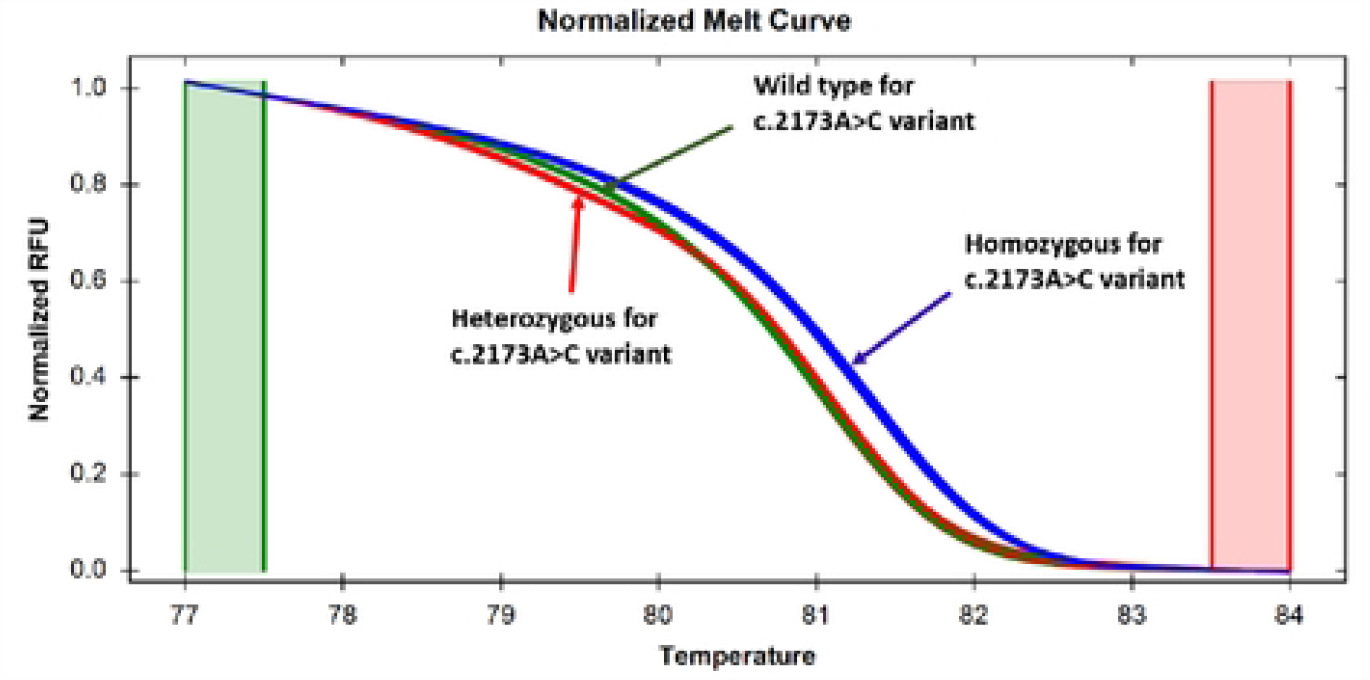
Normalized melt curves for specimens targeting the c.2173A>C variant in exon-12. Discernable changes in normalized melt curves were showing that the specimens with homozygous and heterozygous states are clearly distinguishable from the wild-type alleles, as manifested by the difference in relative fluorescence unit.

**Fig 6.**
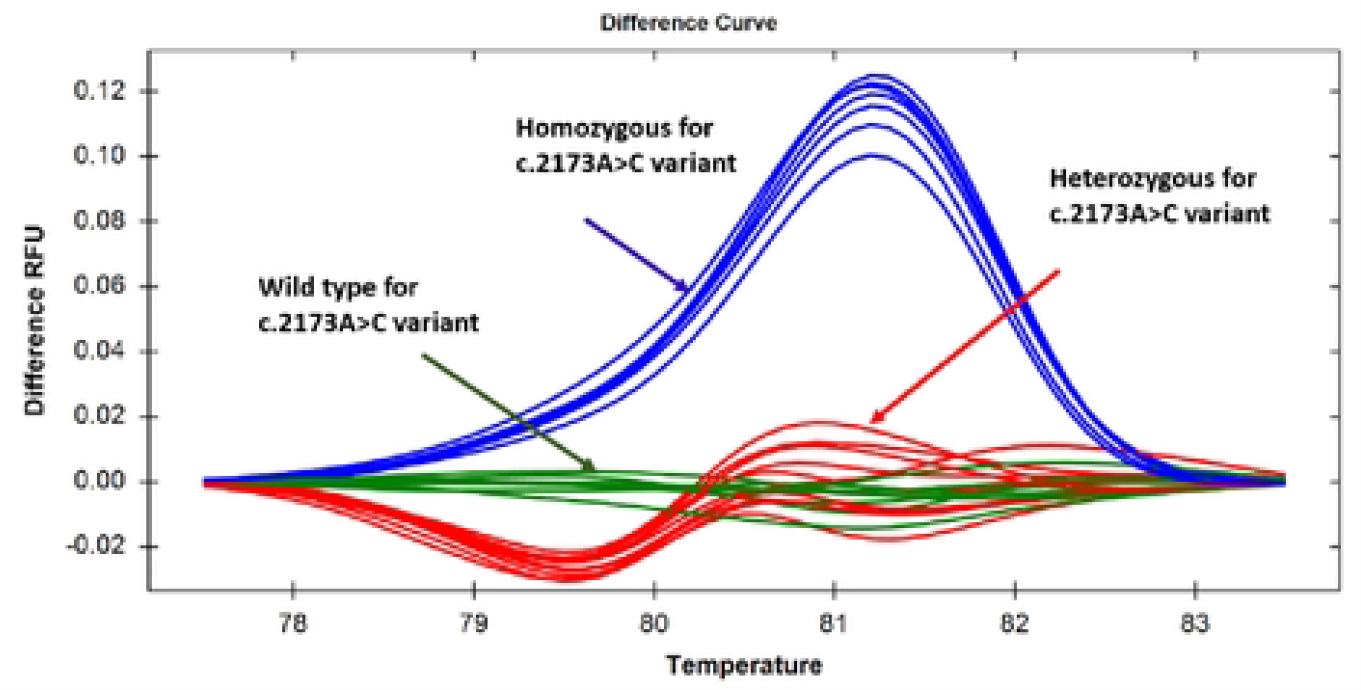
Differential curves for specimens targeting the c.2173A>C variant in exon-12. Discernable changes in difference curves showing specimens with homozygous and heterozygous states are clearly distinguishable from the wild type, as manifested by the difference in relative fluorescence unit.

## 4. Discussion

Congenital hypothyroidism (CH) is the most common cause of intellectual disabilities in children [27]. If early detection of CH is performed and treatment is initiated within 28 days of birth, clinical complications can be reversed by treatment with Levothyroxine, which is very easy to administer and affordable. Although 11 genes have been reported to be responsible for all CH cases with genetic aetiology, only seven genes are responsible for thyroid dyshormonogenesis, and published data have suggested that one of the major genes for thyroid dyshomonogenesis is thyroid peroxidase (TPO), and its mutations are inherited in an autosomal recessive manner to cause the disease [13, 28, 29]. TPO enzyme catalyzes the iodine oxidation process in the thyroid hormone synthesis pathway [30]. To date, approximately 60 mutations in the *TPO* gene have been reported in a total of 17 exons in the *TPO* gene [14, 31-33]. Global publications on the *TPO* gene in hypothyroid patients demonstrated that most of the mutations were confined between exon 7 and exon 14, and very few mutations had been identified outside this region [32][34]. Although the mutations had been detected as homozygous or heterozygous states, our study confirmed that both alleles of the *TPO* gene of all 36 hypothyroid patients had been affected by mutations, further confirming the recessive pattern of this disease. The identified nonsynonymous mutations had previously been reported to be pathogenic or disease-causing mutations [12, 18].

A genetic study investigated that mutation c.1117G>T and c.2173A>C showed a non-enzymatic reaction rate, and mutation c.1193G>C showed a slightly reduced enzymatic reaction rate compared to the wild-type TPO protein [34]. Our previous study identified four common mutations in the hotspot region from exon 8 to exon 12 in the TPO gene and studied their effect on the 3D structure of the TPO protein [19]. Since we found these common mutations in Bangladeshi patients, we aimed to establish an alternative method of Sanger sequencing to screen the patients. In Bangladesh, there is very little information about newborn screening and the genetic aetiology of CH. High-resolution melting (HRM) methodology represents a significant advancement in mutation detection over the years. The HRM method has been established for the detection of variants of the beta-globin gene in thalassemia patients and G6PD deficiency in Bangladesh [24, 35].

The genetic study is important to investigate the cause of CH. There are some screening methods for the diagnosis of CH, such as measurement of serum/blood TSH, T3, and T4. However, these approaches can only confirm the CH cases but not the actual aetiology. That is, the conventional screening method for CH cannot say whether it is acquired or genetic. If the actual aetiology is known, the duration of treatment can be defined based on the causes. If it is due to a genetic cause, the patients could be enrolled for levothyroxine treatment for their whole life. On the other hand, treatment should be continued for the first three years of life for an acquired cause [20]. So, the treatment strategy will be different for CH cases with genetic aetiology and other reasons for CH with dyshormonogenesis. If the genetic basis of CH is defined in the country, carrier screening is possible to target the underlying genetic cause. If the parents are found to be carriers of CH involving the TPO gene, their children or newborns could be screened, and appropriate measures can be taken, such as early initiation of treatment, which would help to prevent mental retardation. Late diagnosis of CH is common in our country, and an initial pilot study suggested that late-diagnosed hypothyroid children had clinical complications even under levothyroxine treatment in Bangladesh. So newborn screening should be a common practice for early CH diagnosis to prevent mental retardation due to late diagnosis.

The present study aimed to establish HRM-targeted mutations c.1117G>T, c.1193G>C, and c.2173A>C commonly found in Bangladeshi patients. To validate the method, we designed primers covering the mutational hotspot, keeping the product size between 65-101 base pairs, which fulfilled the requirement of the HRM strategy [36]. To establish HRM, the samples with heterozygous, homozygous, and wild-type alleles were subjected to an experiment. For the first set of primers targeted the mutation c.1117G>T, both homozygous and heterozygous states were clearly distinguishable from wild-type alleles. However, c.1193G>C mutation was much more difficult to differentiate due to the formation of a similar number of hydrogen bonds for G>C substitution, and thus similar level bond energy was involved for both the wild type and mutant variants. To overcome these difficulties, the optimum concentration of MgCl_2_ was determined to be 8 mM for detection of a single G>C point mutation by HRM because 8 mM MgCl_2_ concentration could clearly distinguish among homozygous, heterozygous, and wild-type alleles. This showed that MgCl_2_ could have an effect on HRM studies to differentiate different states of mutation of G/C alleles. For the c.1193G>C mutation, the melt curve showed almost similar patterns among the samples with the wild-type allele and also samples with homozygous and heterozygous alleles. However, the temperature-shifted curve could clearly differentiate all the states. Different studies demonstrated that the HRM method could not distinguish purine to pyrimidine nucleotide substitution, such as A to T or G to C substitution, due to the same melting temperature [36-38]. For the third mutation, c.2173A>C, Adenine nucleotide was substituted by Cytosine nucleotide, and due to the difference in bond energy between Purine and pyrimidine group, the melting temperature was shifted for both heterozygous and homozygous states compared to the wild type state. The temperature-shifted pattern was differentiated in such a manner that the wild type had a lower Tm pattern compared to the heterozygous and homozygous states.

Although Sanger sequencing is the gold standard for mutation detection, HRM can be used as a fast and less expensive supplemental approach with 100% sensitivity and specificity for screening and detection of mutations in the TPO gene in Bangladeshi patients. Since TPO gene mutation is inherited in an autosomal recessive manner to cause dyshormonogenesis, this HRM method can also investigate its carrier state.

## 5. Conclusion

High-resolution melt curve analysis could be an alternative approach for screening common mutations in the *TPO* gene in Bangladeshi patients with thyroid dyshormonogenesis so that complications of late-diagnosed patients can be prevented by early screening and initiation of treatment in a different strategy.

## Data Availability

All relevant data are within the manuscript and its Supporting Information files.

## Acknowledgements

The authors are thankful to University Grants Commission (UGC) of Bangladesh for its generous support.

## Supporting information

**S1 Table. Sequenced samples that were used for the HRM method setup**.

**S2 Table. Unknown samples used for HRM method validation**.

## References

1. Rastogi MV, LaFranchi SH. Congenital hypothyroidism. Orphanet journal of rare diseases. 2010;5(1):17.

2. Razavi Z, Mohammadi L. Permanent and transient congenital hypothyroidism in Hamadan West Province of Iran. International journal of endocrinology and metabolism. 2016;14(4).

3. Grüters A, Krude H. Detection and treatment of congenital hypothyroidism. Nature Reviews Endocrinology. 2012;8(2):104.

4. LaFranchi SH, Austin J. How should we be treating children with congenital hypothyroidism? Journal of Pediatric Endocrinology and Metabolism. 2007;20(5):559–78.

5. Klett M. Epidemiology of congenital hypothyroidism. Experimental and Clinical Endocrinology & Diabetes. 1997;105(S 04):19–23.

6. Harris KB, Pass KA. Increase in congenital hypothyroidism in New York State and in the United States. Molecular genetics and metabolism. 2007;91(3):268–77.

7. Deladoëy J, Bélanger N, Van Vliet G. Random variability in congenital hypothyroidism from thyroid dysgenesis over 16 years in Quebec. The Journal of Clinical Endocrinology & Metabolism. 2007;92(8):3158–61.

8. Olney RS, Grosse SD, Vogt RF. Prevalence of congenital hypothyroidism—current trends and future directions: workshop summary. Pediatrics. 2010;125(Supplement 2):S31–S6.

9. Hasan M, Nahar N, Ahmed A, Moslem F. Screening for congenital hypothyroidism-a new era in Bangladesh. SOUTHEAST ASIAN JOURNAL OF TROPICAL MEDICINE AND PUBLIC HEALTH. 2004;34:162–4.

10. Park S, Chatterjee V. Genetics of congenital hypothyroidism. Journal of medical genetics. 2005;42(5):379–89.

11. Grasberger H, Refetoff S. Genetic causes of congenital hypothyroidism due to dyshormonogenesis. Current opinion in pediatrics. 2011;23(4):421.

12. Avbelj M, Tahirovic H, Debeljak M, Kusekova M, Toromanovic A, Krzisnik C, et al. High prevalence of thyroid peroxidase gene mutations in patients with thyroid dyshormonogenesis. European journal of endocrinology. 2007;156(5):511–9.

13. Rivolta CM, Esperante SA, Gruñeiro-Papendieck L, Chiesa A, Moya CM, Domené S, et al. Five novel inactivating mutations in the thyroid peroxidase gene responsible for congenital goiter and iodide organification defect. Human mutation. 2003;22(3):259-.

14. Bakker B, Bikker H, Vulsma T, de Randamie JS, Wiedijk BM, de Vijlder JJ. Two decades of screening for congenital hypothyroidism in The Netherlands: TPO gene mutations in total iodide organification defects (an update). The Journal of Clinical Endocrinology & Metabolism. 2000;85(10):3708–12.

15. Kotani T, Umeki K, Kawano Ji, Suganuma T, Hishinuma A, Ieiri T, et al. Partial iodide organification defect caused by a novel mutation of the thyroid peroxidase gene in three siblings. Clinical endocrinology. 2003;59(2):198–206.

16. Rodrigues C, Jorge P, Soares JP, Santos I, Salomao R, Madeira M, et al. Mutation screening of the thyroid peroxidase gene in a cohort of 55 Portuguese patients with congenital hypothyroidism. European Journal of Endocrinology. 2005;152(2):193–8.

17. Wu J, Shu S, Yang C, Lee C, Tsai F. Mutation analysis of thyroid peroxidase gene in Chinese patients with total iodide organification defect: identification of five novel mutations. Journal of Endocrinology. 2002;172(3):627–35.

18. Balmiki N, Bankura B, Guria S, Das TK, Pattanayak AK, Sinha A, et al. Genetic analysis of thyroid peroxidase (TPO) gene in patients whose hypothyroidism was found in adulthood in West Bengal, India. Endocrine journal. 2014;61(3):289–96.

19. Begum M, Islam MT, Hossain SR, Bhuyan GS, Halim MA, Shahriar I, et al. Mutation spectrum in TPO gene of Bangladeshi patients with thyroid dyshormonogenesis and analysis of the effects of different mutations on the structural features and functions of TPO protein through in silico approach. 2019;2019.

20. Büyükgebiz A. Newborn screening for congenital hypothyroidism. Journal of Pediatric Endocrinology and Metabolism. 2006;19(11):1291–8.

21. Begum MN, Mahtarin R, Ahmed S, Shahriar I, Hossain SR, Mia MW, et al. Investigation of the impact of nonsynonymous mutations on thyroid peroxidase dimer. 2023;18(9):e0291386.

22. Begum MN, Mahtarin R, Islam MT, Ahmed S, Konika TK, Mannoor K, et al. Molecular investigation of TSHR gene in Bangladeshi congenital hypothyroid patients. 2023;18(8):e0282553.

23. Yan J-b, Xu H-p, Xiong C, Ren Z-r, Tian G-l, Zeng F, et al. Rapid and reliable detection of glucose-6-phosphate dehydrogenase (G6PD) gene mutations in Han Chinese using high-resolution melting analysis. The Journal of Molecular Diagnostics. 2010;12(3):305–11.

24. Islam MT, Sarkar SK, Sultana N, Begum MN, Bhuyan GS, Talukder S, et al. High resolution melting curve analysis targeting the HBB gene mutational hot-spot offers a reliable screening approach for all common as well as most of the rare beta-globin gene mutations in Bangladesh. BMC genetics. 2018;19(1):1.

25. Bono C, Nuzzo D, Albeggiani G, Zizzo C, Francofonte D, Iemolo F, et al. Genetic screening of Fabry patients with EcoTILLING and HRM technology. BMC research notes. 2011;4(1):323.

26. Bataille S, Berland Y, Fontes M, Burtey S. High Resolution Melt analysis for mutation screening in PKD1 and PKD2. BMC nephrology. 2011;12(1):57.

27. Grosse SD, Van Vliet G. Prevention of intellectual disability through screening for congenital hypothyroidism: how much and at what level? Archives of disease in childhood. 2011:archdischild190280.

28. Pannain S, Weiss RE, Jackson CE, Dian D, Beck JC, Sheffield VC, et al. Two different mutations in the thyroid peroxidase gene of a large inbred Amish kindred: power and limits of homozygosity mapping. The Journal of Clinical Endocrinology & Metabolism. 1999;84(3):1061–71.

29. Fugazzola L, Mannavola D, Vigone MC, Cirello V, Weber G, Beck-Peccoz P, et al. Total iodide organification defect: clinical and molecular characterization of an Italian family. Thyroid. 2005;15(9):1085–8.

30. Taurog A, Dorris M, Doerge DR. Evidence for a radical mechanism in peroxidase-catalyzed coupling. I. Steady-state experiments with various peroxidases. Archives of biochemistry and biophysics. 1994;315(1):82–9.

31. Belforte FS, Miras MB, Olcese MC, Sobrero G, Testa G, Muñoz L, et al. Congenital goitrous hypothyroidism: mutation analysis in the thyroid peroxidase gene. Clinical endocrinology. 2012;76(4):568–76.

32. Bikker H, Vulsma T, Baas F, de Vijlder JJ. Identification of five novel inactivating mutations in the human thyroid peroxidase gene by denaturing gradient gel electrophoresis. Human Mutation. 1995;6(1):9–16.

33. Kimura S, Hong YS, Kotani T, Ohtaki S, Kikkawa F. Structure of the human thyroid peroxidase gene: comparison and relationship to the human myeloperoxidase gene. Biochemistry. 1989;28(10):4481–9.

34. Guria S, Bankura B, Balmiki N, Pattanayak AK, Das TK, Sinha A, et al. Functional analysis of thyroid peroxidase gene mutations detected in patients with thyroid dyshormonogenesis. International journal of endocrinology. 2014;2014.

35. Islam MT, Sarker SK, Talukder S, Bhuyan GS, Rahat A, Islam NN, et al. High resolution melting curve analysis enables rapid and reliable detection of G6PD variants in heterozygous females. BMC genetics. 2018;19(1):58.

36. Słomka M, Sobalska-Kwapis M, Wachulec M, Bartosz G, Strapagiel D. High Resolution Melting (HRM) for High-Throughput Genotyping—Limitations and Caveats in Practical Case Studies. International journal of molecular sciences. 2017;18(11):2316.

